# Accuracy of menu calorie labelling in the England out-of-home food sector during 2024: Assessment of a national food policy

**DOI:** 10.1101/2024.10.24.24316051

**Authors:** Amy Finlay, Andrew Jones, Paula Thorp, I Gusti Ngurah Edi Putra, Megan Polden, Jean Adams, Jane Brealey, Eric Robinson

## Abstract

Mandatory calorie labelling was introduced in out-of-home (OOH) food sector outlets during 2022 in England. Previous research in North America has found labelled energy content can be underestimated for packaged and quick-serve foods, but no study has evaluated the accuracy of out-of-home food sector menu calorie labelling in response to the mandatory policy introduced in England. N=295 menu items from a range of outlet types (e.g. cafes, pubs, restaurants) and menu categories (e.g., starters and sides, main, dessert) were sampled. Bomb calorimetry was used to quantify energy content and the reported energy content on menus was recorded. Consistency of measured energy was assessed by sampling the same items across outlets of the same business (N=50 menu items). Differences between reported and measured energy content were tested through Wilcoxon Signed Rank tests, and a linear model examined correlates of the difference. Mean measured kilocalories (kcal) were significantly lower than reported kcal (−16.70kcal (±149.19), V=16920, p<.01, r=0.182). However, both over- and under-estimation of measured energy content was common and the averaged absolute percentage difference between reported and measured values was 21% (±29%). Discrepancy between measured and reported energy content was more common in some outlet types (pubs) and reported energy content was substantially different (>20%) to measured energy content for 35% of sampled menu items. There were significant inaccuracies in reported energy content of calorie labelled menu items in English food outlets subject to mandatory calorie labelling and this appears to be caused by both over- and under-estimation of reported energy content.

**Highlights:**

- Energy content measured by bomb calorimetry was significantly higher than on menus.
- Both under and over estimation of food energy content were frequently observed.
- Measured energy content was consistent across chain outlets in different locations.

## 1. Introduction

In April 2022, calorie labelling on food menus was made mandatory in England for all businesses with over 250 employees. This policy was specific to the out-of-home (OOH) food sector, and applied to all non-prepacked foods and drinks made for immediate consumption(1). Consumption of OOH food is associated with poorer dietary quality and increased daily energy intake(2, 3). Due to these concerns, the government in England introduced mandatory calorie labelling to promote informed food choices and healthier eating behaviour in the OOH food sector through consumers substituting high energy foods for lower energy foods, reformulation of foods by the food industry to reduce energy content and/or introduction of new lower energy foods to menus(4). Calorie labelling policies have been implemented outside of the UK, specifically in parts of Australia(5), Canada(6) and in the US nationally(7). Evidence is mixed on the impact that calorie labelling has on consumer behaviour(8, 9), but studies tend to suggest that calorie labelling results in reductions to energy content of menu items(10, 11).

Implementation guidance for the 2022 mandatory calorie labelling policy in England(4) states that businesses can calculate the energy content of menu items by averaging values based on manufacturers analysis, known or average values of ingredients used, or established and accepted food databases. The energy content of menu items must be displayed alongside the portion size to which it relates, and a statement that ‘adults need around 2000 kcal a day’ at any point of choice for the consumer. A 20% discretion between the reported and calculated energy content is permitted, but guidance acknowledges that accurate testing of energy content may not be viable for local enforcement officers(4). Regarding enforcement, this guidance states that “Local authorities have discretion in how they enforce the Regulations”(4). Specific roles of the enforcement officer include checking calorie labels are present and that the methods used to estimate energy content are appropriate(4). However, there is little guidance for enforcement officers in assessing the accuracy of calorie labelling and no explicit instruction or suggestion that accuracy of labelling of menu items should be tested.

In the US, businesses appeared initially to have largely complied with the mandatory provision of calorie labelling, with 79% of businesses fully or partially implementing labels in advance of policy enforcement(12) and similarly in the UK, 80% of businesses provided calorie labelling at any point of choice 6 months post-implementation, compared to only 21% who provided this information prior to implementation(13). However, more recent evidence in the US shows that large chains may not be reporting calorie information across all required formats(7). The food industry has a history of inadequate compliance and exploitation of loopholes with government policy(14-16). It is likely that this is due to the lack of accountability by the food industry surrounding implemented policy and the lack of resources for compliance monitoring(17).

There is evidence that nutritional labels can be prone to some degree of inaccuracy for packaged food products. One study assessed the accuracy of nutritional labelling of popular snacks in the US and found that the energy content of snacks was on average higher than reported on labels(18). However, for most items (96%), reported energy was within 20% of measured energy content. In Canada, over 1000 items from supermarkets, bakeries and restaurants were tested for their nutritional content(19). For the tested foods, sodium and energy were consistently under-reported. For 14% of foods, the measured energy content exceeded label values by more than 20%. A study conducted in the US assessed the accuracy of nutrition labels for low-energy (<500kcal) restaurant and frozen meals(20). For frozen meals, measured energy content was on average 8% higher than reported and for restaurant meals energy content was on average 18% higher than reported. Several restaurant foods contained up to twice the reported energy content. Similarly, a US study of restaurant menu items in 2011 found that although average measured vs. reported energy content across menu items was similar, 19% of individual menu items had a measured energy content of ≥100kcal per portion more than reported(21).

No research has to date assessed the accuracy of calorie labels in OOH outlets following the implementation of the mandatory calorie labelling policy in England. As customers are encouraged to pay attention to the nutritional labelling of foods(1), and it is expected that the provided information will impact behaviour, inaccurate nutritional labelling may hinder individual efforts to eat healthier. For example, if consumers use calorie labels to factor foods into their daily energy allowance, then underestimation of calories on food menus would lead to consumers unknowingly consuming excess energy. Additionally, if consumers identify potential inaccuracies in labelling themselves, then they may lose trust and stop using calorie information in the OOH food sector. Furthermore, some research suggests there is little or no enforcement of the policies requirement of providing accurate calorie information(22). Therefore, in the present study we examined the accuracy of menu calorie labelling in OOH sector outlets subject to the 2022 mandatory calorie labelling law in England.

## 2. Materials and methods

This study was pre-registered on the Open Science Framework https://osf.io/8tfu4/.

### 2.1. Outlet selection

Outlets in two local authorities (LAs) in England were sampled to ensure findings were not area specific. Liverpool (North of England) and Milton Keynes (South of England) were selected to ensure mixed geographical coverage and representation of different quintiles of deprivation (Index of Multiple Deprivation (IMD) used at the LA level). LA IMD quintiles (1-5) were used with IMD1 reflecting the most deprived areas and IMD5 reflecting the least deprived. LAs selected represent quintile 1 (most deprived – Liverpool) and quintile 3 (medium deprivation – Milton Keynes) however outlets in both Las were located across quintiles 1-5 as measured at the Lower Super Output Area.

For previous research that evaluated the impact of the calorie labelling legislation on consumer and business behaviour(23), the Inter-Department Business Register (IDBR) was used to identify eligible businesses likely to be subject to the mandatory calorie labelling policy within LAs of interest (data sampled in June 2021, list produced in Autumn 2020). The IDBR is a list of all UK businesses, which includes their core characteristics, number of employees and principal activities defined using the Standard Industrial Classification. We identified Standard Industrial Classification codes likely to include businesses serving food (for the full list of Standard Industrial Classification codes used see Supplementary Material 1) and then identified large businesses with 250 or more employees. Individual businesses often have multiple outlets (e.g. chain restaurants), so individual outlets belonging to each identified large business within the LAs of interest were identified using Ordnance Survey Points of Interest data from September 2020.

From this database, we randomly selected (using the RAND function in excel) N=6 unique outlets from each of four main outlet categories: café, restaurant, fast food, pubs. If for any outlet category there were not 6 unique outlets (i.e., outlets from different chains), additional outlets classed as ‘sport & entertainment’ were sampled. We randomly selected additional outlets from the full database until N=25 unique outlets were selected for each LA. Duplicates of businesses across the two LAs (e.g. where outlets of the same large business were sampled in both LAs) were permitted. Once all outlets had been selected, n=5 of the selected outlets (20%) in each LA were randomly selected and matched to an additional corresponding outlet from the same business in the other LA where the same items would be sampled. This subsample of matched outlets allowed for the assessment of consistency in energy content across outlet chains in different LAs. Therefore, in total we sampled menu items from a total of 60 outlets (30 from each LA).

### 2.2. Menu item selection

Prior to data collection, online menus for each outlet were used to identify the items on offer in each of five food categories. For each outlet, one menu item was selected for sampling in each of the following categories:

- Starter/side or prepared drink if starter/sides not readily available (randomly selected)
- Main meal (randomly selected)
- Dessert (randomly selected)
- Most popular menu item (determined by asking serving staff)
- Menu item that that was potentially inaccurate (determined by a research staff member and trained nutritionist assessing the full menu)

In instances where menu categories were not distinct (e.g. starter – main – dessert – side dish), two researchers individually considered the categories provided by outlets and re-categorised items into the above groups. Any differences were resolved through discussion. If it was anticipated that the categories outlined above would not encompass the majority of items available in outlets (e.g., coffee shops), then prepared drinks were sampled. This was the case for n=7 outlets from n=3 unique businesses.

Items in each category were numbered and a random number generator was used to select items for sampling. For the items sampled under the category of potentially inaccurate, two researchers considered the composition of menu items and whether the reported energy content looked inaccurate using menu information alone (either too high or too low). If there were no items that appeared to be inaccurate, the researchers considered instances where potential variability in serving size made by staff (e.g., inconsistency in the serving size of a dish of pasta) could result in differences in energy content from reported. Once each researcher had determined a potentially inaccurate item for each outlet, a final item was decided upon through discussion. In each outlet, the researcher also asked the serving staff what the most popular item on the menu was, and this item was sampled. If the most popular menu item or potentially inaccurate item from an outlet was a drink, this was eligible for sampling.

For instances where menu items required customisation (e.g. choosing a side to go with a dish), we randomly selected from all available customisation options. In total, 300 items from food menus were collected from 60 outlets across two LAs in England. For sample size and power analysis information see Supplementary Material 2.

### 2.3. Energy content of sampled items and procedure

The researcher responsible for data collection recorded the energy content reported on menus for each sampled menu item on the day of sampling. Samples were collected (Monday-Thursday) between 11am-7pm during April-May 2024. For each outlet, the researcher responsible for data collection ordered food to dine in. All sampled items were individually weighed and packaged in the restaurant. Calibration weights were used to ensure scales were accurate. All items were sent via a courier to an external laboratory accredited by the UK Accreditation Service (SGS Cambridge) for nutritional analysis through bomb calorimetry and energy (kcal) data were analysed.

### 2.4. Analysis

### 2.4.1. Primary analyses

For all analyses, the potentially inaccurate items were considered separately from all other items tested as we anticipated their inclusion may overestimate average difference between measured and reported energy content. For the main analyses, one item was missing due to lab error (spoiled food due to storage error), and three items were excluded from analysis due to implausibility of energy values. This resulted in n=236 samples. Similarly, for potentially inaccurate items, one menu item was missing due to lab error, resulting in n=59 samples.

Throughout the results, relative differences are calculated as *measured kcal – reported kcal* for both mean and percentage difference. Absolute differences are calculated as the mean or percentage difference from measured kcal regardless of the direction of the difference, calculated using the ‘abs()’ function in R.

Absolute percentage differences were used to explore whether differences between measured and reported energy content differed from 0% (perfect accuracy) and 20% (permitted inaccuracy). As data were not normally distributed, one-sample Wilcoxon signed rank tests were conducted against 0 and 20% to examine if the observed absolute difference values were significantly different from 0% and 20%.

We also conducted Wilcoxon signed rank tests to assess whether there was a significant difference between reported energy and lab-measured energy content (relative difference).

### 2.4.2. Secondary analyses

For the subsample of restaurants where a corresponding outlet from each LA was tested, resulting in n=50 matched pairs, we examined whether the measured energy content differed between the two locations using a paired Wilcoxon signed rank test.

We created a new variable [measured kcal – reported kcal] and fitted a linear regression model to examine potential predictors (e.g., outlet characteristics) of the relative difference between measured and reported energy. We explored outlet type, menu category, item energy content, IMD score for outlet and LA. If any predictor variables were significant, we conducted subgroup analyses to explore differences. We also used this method to explore predictors of absolute mean percentage difference.

Results for primary analyses were considered significant at p <.05. To account for multiple comparisons, results for secondary and any exploratory analyses were considered significant at p <.01. For explanation of deviation to the pre-registered analysis plan see Supplementary Material 3.

## 3. Results

A total of n=295 menu items were sampled from N=60 outlets across two LAs in England. Figure 1 displays the mean reported and measured energy content for each category of item. Menu items were categorised by outlet type (pubs, cafes, fast food, restaurants and entertainment) and menu category (starter/side, main, dessert, popular item, potentially inaccurate item and drinks). Mean energy and percentage differences between reported and measured energy for each category are reported in Table 1. Overall, the mean relative difference between reported and measured energy (measured-reported) was –16.70 kcal (±149.19) and -8.75% (±35.27%) expressed as a relative percentage of the menu item energy content. This was 98.76 kcal ±112.88 and 21.30% (±29.45%) when the absolute energy and percentage difference were examined (i.e. mean difference regardless of direction of difference).

**Table 1:** Difference between reported and measured energy (kcal) for the different categories.

|  | Mean kcal content | Relative mean difference (measured-reported) (kcal)* | Relative percentage difference** | Absolute mean difference*** | Absolute percentage difference*** | Percent outside 20% leeway |
| --- | --- | --- | --- | --- | --- | --- |
| Overall (N=236) | 560.40 (329.11) | -16.70 (149.19) | -8.75% (35.27%) | 98.76 (112.88) | 21.30% (29.45%) | 34.75% |
| **Outlet category** |  |  |  |  |  |  |
| Cafes (N=44) | 313.03 (158.74) | -2.60 (48.96) | -3.44% (20.52%) | 36.29 (32.51) | 15.02% (14.30%) | 29.55% |
| Pubs (N=51) | 674.00 (338.37) | -51.00 (154.94) | -20.08% (50.54%) | 126.26 (101.97) | 29.78% (45.46%) | 45.10% |
| Fast food (N=51) | 523.75 (323.79) | -8.62 (134.77) | -4.97% (27.13%) | 82.89 (105.98) | 17.73% (21.01%) | 29.41% |
| Restaurants (N=76) | 660.67 (318.30) | -21.97 (175.63) | -8.71% (31.73%) | 123.30 (126.22) | 21.38% (24.96%) | 35.53% |
| Entertainment (N=14) | 552.79 (330.88) | 63.14 (204.14) | 1.91% (43.10%) | 119.50 (174.89) | 22.71% (35.88%) | 28.57% |
| **Item category** |  |  |  |  |  |  |
| Starter (N=52) | 342.85 (220.37) | -17.61 (127.99) | -19.42 (55.76) | 84.88 (96.71) | 33.44% (48.60%) | 48.08% |
| Main (N=59) | 763.98 (360.26) | 6.10 (184.10) | -3.77 (22.36) | 122.21 (136.88) | 16.15% (15.84%) | 28.81% |
| Dessert (N=60) | 520.73 (190.61) | -31.23 (132.03) | -6.66 (28.66) | 85.35 (104.95) | 17.77% (23.34%) | 23.33% |
| Popular (N=57) | 658.03 (330.78) | -26.45 (154.66) | -7.06 (27.33) | 110.64 (110.33) | 18.75% (20.95%) | 35.09% |
| Drink (N=8) | 144.18 (84.81) | -0.57 (38.06) | -3.74 (30.19) | 31.86 (16.99) | 25.00% (15.00%) | 75.00% |
| Potentially inaccurate (N=59) | 738.16 (378.56) | -35.45 (169.36) | -9.93% (31.56%) | 121.96 (121.78) | 20.88% (25.64%) | 35.59% |
| **Local authority** |  |  |  |  |  |  |
| Liverpool (N=119) | 538.15 (298.88) | -27.03 (130.10) | -8.93% (29.63%) | 87.51 (99.70) | 19.62% (23.91%) | 34.45% |
| Milton Keynes (N=117) | 587.77 (354.39) | -6.19 (166.28) | -8.56% (40.33%) | 110.20<br>(124.26) | 23.01% (34.19%) | 33.33% |

\*Negative values indicate that reported kcal were greater than measured
\*\*Percentage difference for reported – measured kcal
\*\*\*the mean difference between reported and measured kcal regardless of direction (- or +) of difference
Numbers in brackets are standard deviations

**Figure 1:**
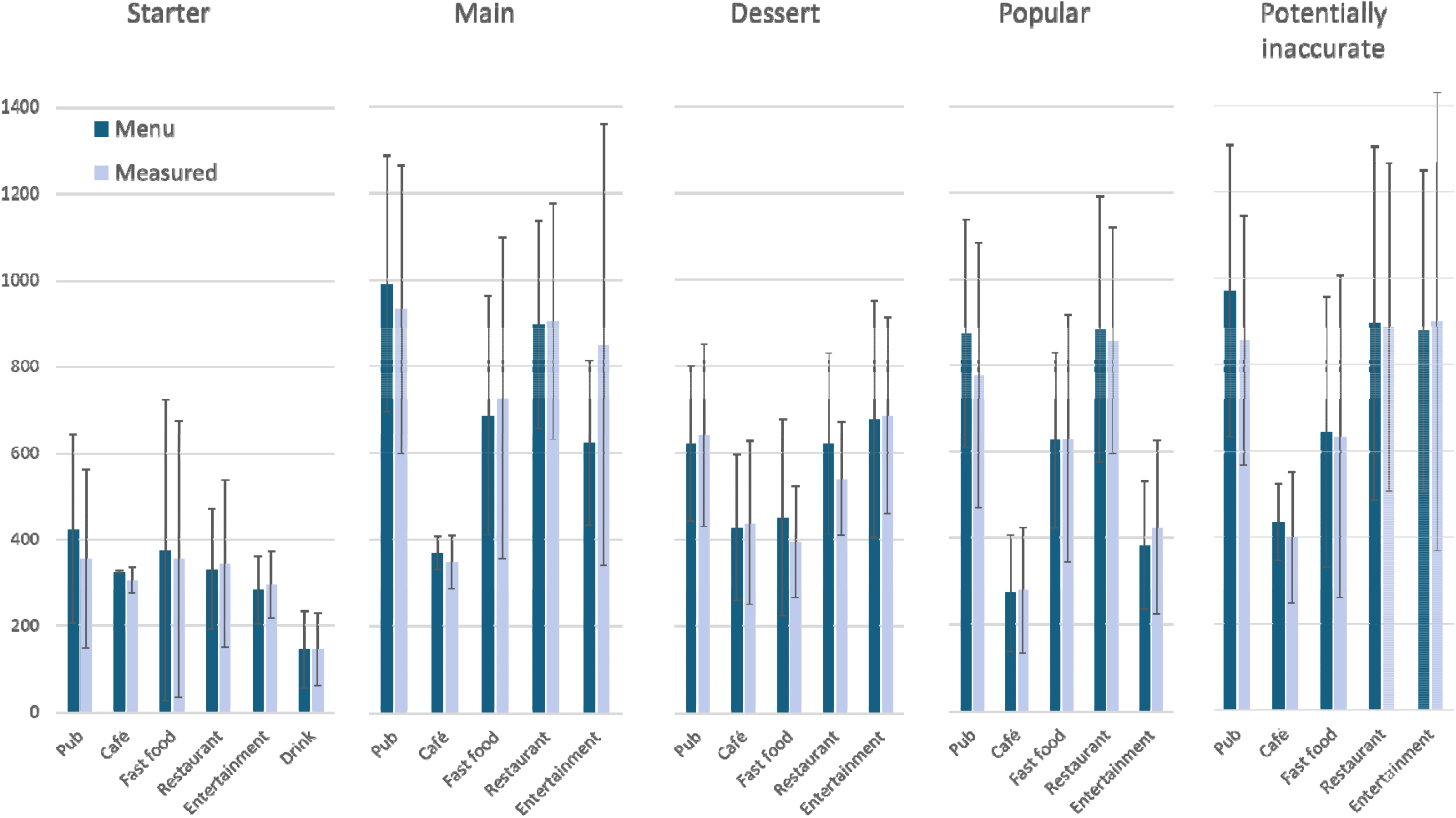
Measured vs reported energy (kcal) split by outlet type and food category. ^*^Error bars represent standard deviation

In the main analyses (all menu items except for those that were potentially inaccurate), 56% of items had a lower measured energy content than reported on menus. For over one third (35%) of items, the energy content reported on menus was outside of the 20% leeway permitted; for 66% of these (23% of total) this was due to reported energy being substantially higher than measured and for 31% of these (11% of total) this was due to reported energy being substantially lower than measured. Findings were similar in the potentially inaccurate items (See supplementary Material 4).

Two Wilcoxon signed rank tests found that the absolute percentage difference between reported and measured energy was significantly greater than 0% (V=27261, p<.001) but not greater than 20% (v=0, p>0.99). However, reported and measured energy content were significantly different from each other, whereby on average energy content reported on menus was significantly greater than measured energy content (V=16920, p<.01, r = 0.182). A boxplot of mean differences is shown in Figure 2. Reported and measured energy were not significantly different for potentially inaccurate items.

**Figure 2:**
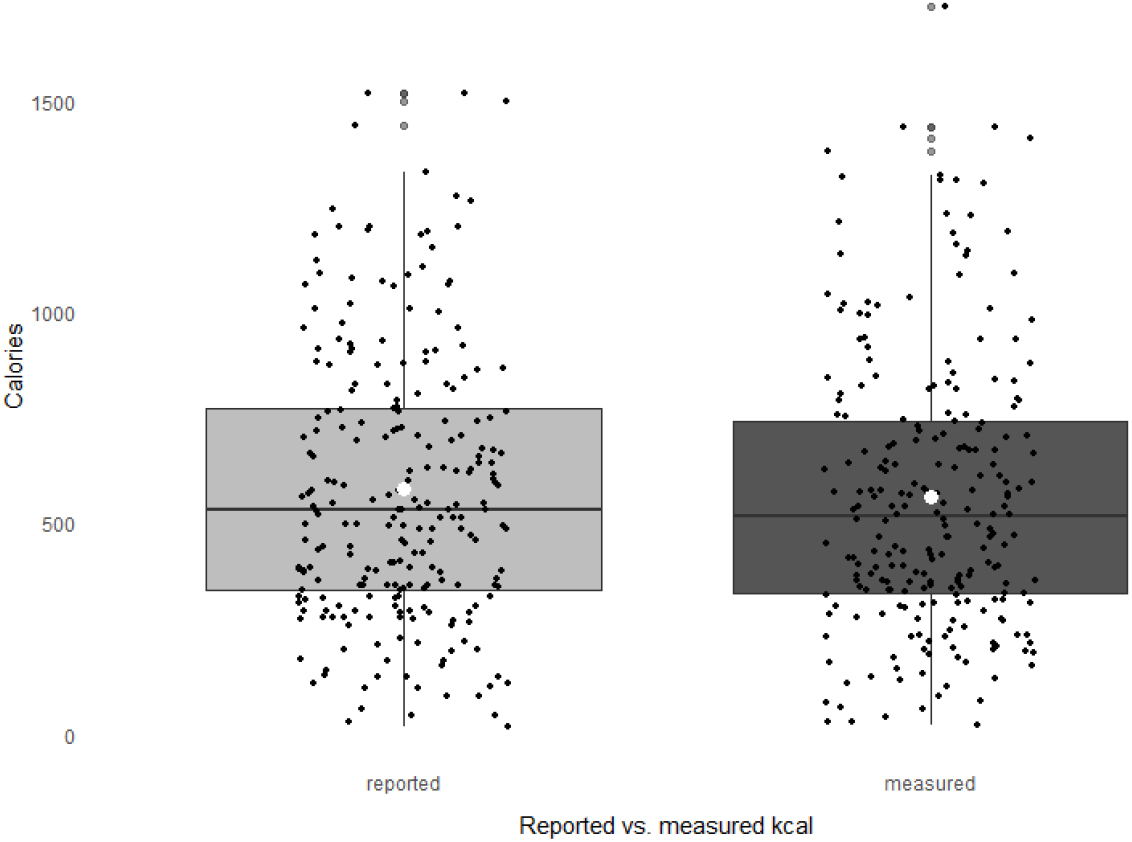
Reported and measured energy content of n=236 items. ^*^white circle indicates the mean

For the subsample of menu items from outlets matched across the two LAs, a Wilcoxon signed rank test found no significant differences in measured energy between matched items (V=583, p=.775). See Figure 3. The mean energy content of these items in Liverpool was 547.39 kcal ± 327.96 and for Milton Keynes this was 549.45 kcal ±312.02. The mean difference (Liverpool – Milton Keynes) was -14.15 kcal, and the relative percentage difference was -7.99%. 73% of items sampled in Milton Keynes were within 20% of the value in Liverpool.

**Figure 3:**
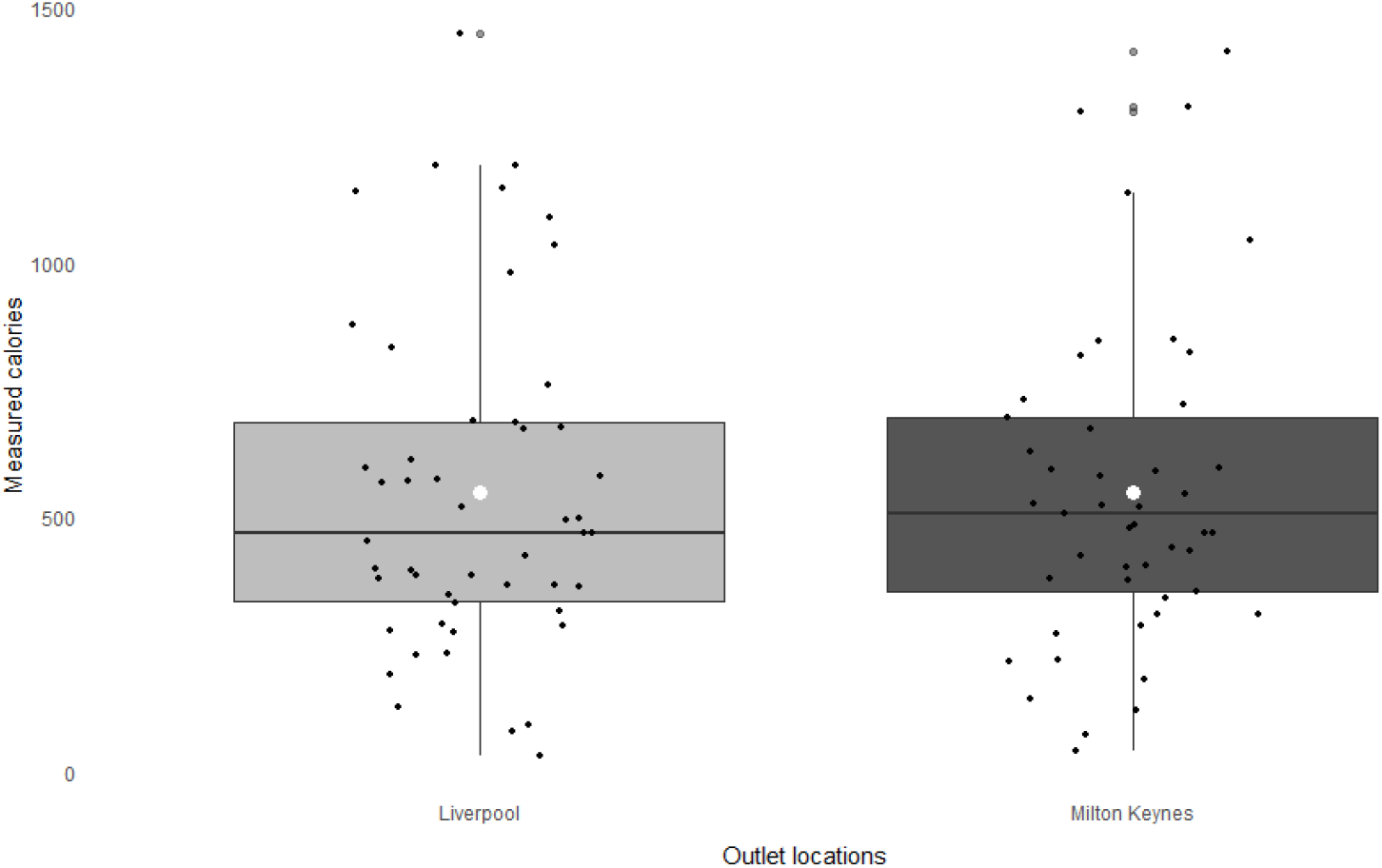
Matched pairs of menu items across the two local authorities. ^*^white circle indicates the mean

A linear model explored predictors of the mean difference between reported and measured energy content (Table 2). Variance Inflation Factor (VIF) for all predictors were below 2, indicating minimal multicollinearity. Items with a higher measured energy content were more likely to have a measured energy higher than reported energy (0.22, 99% CI 0.12 to 0.32, p<.001) whereby every measured 1kcal increase, was associated with a difference of 0.26kcal. Main effects were also observed for outlet type and item category, whereby pubs and restaurants (vs. cafes), and popular dishes (vs. starters/sides) had a greater difference between reported and measured energy. There were no significant predictors of difference between measured and reported energy for potentially inaccurate items (full results in Supplementary Material 4). We also explored predictors of absolute mean percentage difference between reported and measured energy content, and results were similar to main analyses (Supplementary Material 5).

**Table 2:** Linear model exploring predictors of mean relative difference between reported and measured energy content (calculated as measured – reported)

|  | Mean kcal difference |  |  |
| --- | --- | --- | --- |
| *Predictors* | *Estimates* | *99% CI* | *p* |
| (Intercept) | -16.87 | -111.00 – 77.27 | 0.642 |
| Pub* | -134.08 | -222.36 – -45.80 | **<0.001** |
| Fast Food* | -61.09 | -144.82 – 22.64 | 0.059 |
| Restaurant* | -99.88 | -185.36 – -14.39 | **0.003** |
| Entertainment* | 8.66 | -111.15 – 128.47 | 0.851 |
| Main** | -79.16 | -161.72 – 3.39 | 0.013 |
| Dessert** | -64.15 | -136.38 – 8.08 | 0.022 |
| Popular** | -88.89 | -166.98 – -10.79 | **0.003** |
| Drink** | -22.60 | -174.43 – 129.23 | 0.699 |
| IMD quintile [2]*** | -5.59 | -84.61 – 73.43 | 0.854 |
| IMD quintile [3] *** | 3.08 | -55.14 – 61.30 | 0.891 |
| IMD quintile [4] *** | 3.75 | -84.58 – 92.08 | 0.912 |
| IMD quintile [5] *** | -54.97 | -246.44 – 136.50 | 0.456 |
| Outlet location [MK]**** | 15.66 | -34.90 – 66.21 | 0.422 |
| Total energy kcal | 0.22 | 0.12 – 0.32 | **<0.001** |
| Observations | 236 |  |  |
| R<sup>2</sup> / R<sup>2</sup> adjusted | 0.181 / 0.129 |  |  |

Reference categories are: \*Cafes, \*\* Starters/sides, \*\*\*IMD quintile 1, \*\*\*\*Liverpool Negative values indicate a decreased relative difference

To explore identified significant predictors, paired samples tests (Wilcoxon signed ranks) were conducted which examined differences between measured vs reported energy for cafes, restaurants and pubs, and the starter and popular item categories. Pubs were the only outlet type where reported and measured energy were significantly different from each other (V=943, p<.01). For pubs, mean reported kcal were 725.00 ±326.89 and mean measured kcal were 674.10 ±338.36. There was not a significant difference between measured and reported energy for the starter or popular menu categories. Mean differences, percentage differences and the proportion of items outside of the 20% leeway for each outlet type and item category are shown in Table 1.

## 4. Discussion

This study of large business owned OOH food outlets subject to the mandatory calorie labelling law in England during 2024 found that averaged across all sampled menu items, the energy content (kcal) reported on OOH outlet menus was significantly greater than measured energy. However, it was also common for reported energy content of menu items to be greater and less than measured energy content. For 35% of menu items, reported energy was over 20% greater or less than measured energy. Pubs had a particularly pronounced difference between reported and measured energy content compared to other outlet types. The measured energy content of the same menu items sampled from different outlets of the same chain were largely similar. Collectively, these findings suggest that reported energy content for significant numbers of menu items in the OOH sector in England differ substantially to measured energy content.

Expressed as a relative calorie value and a percentage of each item’s measured energy content, average reported energy of items was 17kcal and 9% greater than measured energy content. However, expressed as an absolute value (size of deviation from measured energy content irrespective of direction), the mean difference between measured and reported energy was 99 kcal and 21%, due to both over- and under-estimation of measured energy content being common (although over-estimation of energy content was observed more frequently). This suggests that on average, calories reported for menu items in the English OOH food sector may differ substantially to their measured energy content. The overall pattern of results differs somewhat to research conducted in the US (18, 20) which identified that measured energy content tended to be greater than reported on labelling and on restaurant menus. This difference in our findings may be a result of differences in types of menu items examined and/or sampling methods. One of these US studies considered OOH foods, but limited investigation to menu items under 500kcal(20), which may also explain differences as our findings would suggest there is less scope for inaccuracy at lower levels of energy content. A study from Canada sampled over 1000 items from grocery stores, bakeries and restaurants and did not limit investigation to lower energy items(19). Similar to the present study, overall, the items tested had a greater mean labelled energy content than mean measured energy content, and there was evidence of both over and under-estimation of reported energy content based on measured energy content.

In the previous North American studies discussed, for the majority of menu items, reported energy content was within the 20% leeway of measured content (70%(18), 86%(19), 59%(20)) and this is comparable to the present study (65%). However, across all of these and the present study, a significant proportion of menu item labels appear to be significantly different from measured values. In England, the calorie labelling legislation outlines several accepted methods to calculate energy content. While laboratory measurement (i.e. bomb calorimetry) is the gold standard of measurement, this is expensive, and we assume an unlikely method in how OOH food sector outlets in England calculate the energy content of their menu items. Instead, outlets may be more likely to be basing estimations on manufacturers analysis, known or average values of ingredients used, or established and accepted food datasets(4). It is therefore likely that the observed error, and perhaps bias to overreporting comes from these indirect methods of estimating energy content.

While five different outlet types were explored, the largest difference between mean reported and measured energy content was observed for pubs. This outlet type had 46% of items outside of the 20% legislation leeway, while all other outlet types had between 30-36% of items outside of the leeway. Observed differences between measured and reported energy may have been greater in pubs due to differences in the use of ingredients which are more prone to fluctuations in water and energy content through cooking. Alternatively, differences between outlet types, and individual outlets may exist if there are differences in how the energy content of menu items is calculated.

When items deemed potentially inaccurate were examined, results were similar to the main analysis. Items in this category were selected based on two researcher’s assessments of all items on a menu with consideration of the ingredients and composition of the dish and the likelihood of inconsistencies in portion sizes made by servers in outlets. As these findings were not largely different from the items in the main analyses, this suggests that reliably identifying likely inaccurately labelled menu items based on menu information alone may be difficult. This highlights the difficulty that enforcers of the policy may have if attempting to assess accuracy of calorie labelling in OOH outlets without expending substantial resources on laboratory measurement. At present there appears to be minimal enforcement training supplied, although how enforcement is monitored is at the discretion of the LA(4). To aid improved accuracy of a menu’s calorie information and enforcement of the policy, laboratory analysis of menu items would preferably determine reported energy content, however this would result in significant financial cost to businesses. Despite this, a one-off or annual cost may be minimal for larger food chains such as those explored in this study. Our results showed that the same menu items from different outlet locations of the same business tended to have similar measured calorie content, so analysis at the chain level is likely representative of all outlets in the chain. However, this should be explored in greater depth, and particularly within pubs, where the greatest inconsistencies in reported vs. measured energy content were observed.

There are a number of limitations of this work that should be considered alongside findings. This study explored a large sample of menu items in the OOH food sector; however, this sample is not representative of all menu items in outlets in England and instead provides a snapshot of the accuracy of calorie labels in the OOH food sector among large businesses.

The analyses relating to matched samples and exploration of potentially inaccurate items can be considered as exploratory only, as the study was not powered for these smaller samples. We used a gold standard measurement of energy content, and this is a strength of the present work. However, we collected one sample per menu item from outlets for our main analyses and future research would benefit from taking multiple samples of the same item. This would improve measurement accuracy for estimated energy content and also allow for examination of consistency of menu item energy content within the same outlet.

## 5. Conclusion

There were significant inaccuracies in reported energy content of calorie labelled menu items in English food outlets subject to mandatory calorie labelling and this appears to be caused by both over- and under-estimation of reported energy content.

## Data Availability

Data will be made available on the Open Science Framework at the time of publication

