## Supplementary Material for "Accuracy of menu calorie labelling in the England out-of-home food sector during 2024: Assessment of a national food policy"

**Supplementary Materials**

**Supplementary Material 1: List of Standard Industrial Classification codes used for sampling**

Large businesses with the following Standard Industrial Classification (SIC) codes (and hence in the following IDBR sections) that are subject to the mandatory calorie policy and thus included in this study:

Section I (accommodation and food service activities)

Within section G:

- SIC 47.11- Retail sale in non-specialised stores with food, beverages or tobacco predominating.
- SIC 47.24- Retail sale of bread, cakes, flour confectionery and sugar confectionery in specialised stores.
- SIC 47.29 Other retail sales of food in specialised stores.

Within section R:

- SIC 91.03-Operation of historical sites and buildings and similar visitor attractions
- SIC 91.04-Botanical and zoological gardens and nature reserve
- SIC 93.11-Operation of sports facilities
- SIC 93.12-Activities of sports clubs
- SIC 93.13-Fitness facilities
- SIC 93.21-Activities of amusement parks and theme parks

Within section J:

- SIC 59.14-Motion picture projection activities

**Supplementary Material 2: Sample size calculation and power analysis**

We intended to sample N=300 items in total^[1]^. Given that the decision was made to analyse items that appeared inaccurate separately from the main sample, we conducted power analyses to confirm the appropriateness of final analytic sample sizes. Using G*Power 3.1.3. we calculated that with a sample size of N=239 menu items, at 80% power and Type 1 error probability of p<.05, we would be able to detect small effect size (i.e., differences between calories reported on menu and actual calorie content) of dz=0.17 through a paired-samples Wilcoxon test. To conduct this test for the subsample of items matched across the two local authorities, we calculated that with a sample size of N=59 at 80% power and Type 1 error probability of p<.01, we would be able to detect a medium effect size (dz=0.34)

For a linear multiple regression (fixed model, R^2^ increase), a sample size of N=239 at 80% power with a type 1 error probability of p=.05 and 5 predictor variables would be able to detect a statistically small sized effect when explaining total variance (f^2^=0.05), suggesting reasonable power for the model to be able to explain some variance in measured vs. reported calories, which we deem acceptable for the exploratory nature of this analysis.

**Supplementary material 3 – deviation from pre-planned analysis in study protocol**

In our pre-registered analysis plan, we stated we would explore using a linear model the predictors of mean difference (i.e. relative difference; measured – reported kcal). We also decided to examine the predictors of absolute mean difference (reported in Supplementary material 5) due to the large observed error in both directions (i.e. both over- and under-estimation), and the possibility that examining relative difference alone could obscure the extent of inaccuracy.

**Supplementary Material 4: Findings for items that appeared inaccurate**

For items that appeared inaccurate a priori, the mean relative difference between reported and measured kcal was -35.45±169.36 and 9.93% (±31.56%). Just over a third (35.59%) of menu items were outside of the 20% leeway permitted; for 25%, this was due to reported menu calories being substantially lower than measured and for 10% this was due to reported calories being substantially greater than measured.

**Supplementary Table 1: Mean difference for items that appeared inaccurate.**

|  | Mean difference (measured-reported) (kcal)* |
| --- | --- |
| **Outlet category** |  |
| Cafes | -37.47 (152.53) |
| Pubs | -114.25 (162.77) |
| Fast food | -11.35 (102.67) |
| Restaurants | -7.52 (185.90) |
| Entertainment | 21.30 (294.17) |
| **Overall item category** |  |
| Likely inaccurate | -35.45 (169.36) |
| **Local authority** |  |
| Liverpool | -11.62 (130.66) |
| Milton Keynes | -60.09 (201.24) |

*Negative values indicate that reported kcal were greater than measured.

Two Wilcoxon signed rank tests found that the absolute percentage difference between reported and measured kcal for items that appeared inaccurate were significantly greater than 0% (V=1653, p<.001) but not 20% (V=0, p>0.99). Reported and measured kcal content were not significantly different from each other (V=1059, p=.190). A boxplot of mean differences is shown in Supplementary Figure 1.

**Supplementary Figure 1: Reported and measured kcal content of items that appeared inaccurate (n=59 items)**
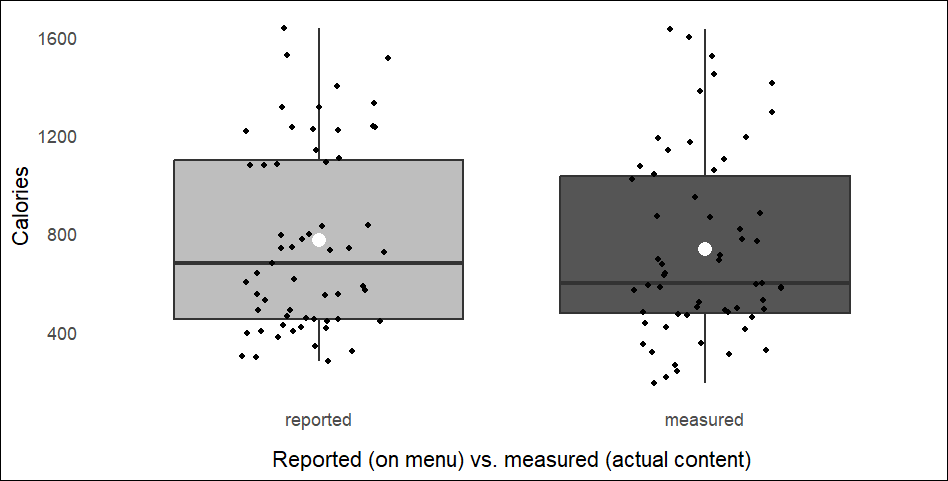

A linear regression model explored predictors of the mean difference between reported and measured kcal for items that appeared inaccurate (Table 2). Here, the VIF for all predictors were below 1.7 so multicollinearity was unlikely to be an issue. The predictors explored were outlet type, item category, outlet IMD, location (local authority) and total energy content. No significant predictors were identified at p<.01.

**Supplementary Table 2: Linear model of predictors of mean difference (measured - reported kcal)**

|  | **Mean difference** | | |
| --- | --- | --- | --- |
| *Predictors* | *Estimates* | *99% CI* | *p* |
| (Intercept) | -43.93 | -241.86 – 154.00 | 0.554 |
| Pubs* | -158.39 | -364.32 - 47.55 | 0.045 |
| Fast food* | -20.78 | -219.14 – 177.59 | 0.780 |
| Restaurant* | -57.59 | -261.32 – 146.15 | 0.452 |
| Entertainment* | -41.84 | -326.62 – 242.94 | 0.695 |
| IMD quintile [2]** | -73.79 | -268.69 – 121.11 | 0.315 |
| IMD quintile [3]** | -8.60 | -151.27 – 134.06 | 0.872 |
| IMD quintile [4]** | -43.23 | -280.36 – 193.90 | 0.627 |
| IMD quintile [5]** | -84.96 | -574.08 – 404.17 | 0.643 |
| Outlet location [MK]*** | -60.20 | -184.07 – 63.67 | 0.199 |
| Total energy kcal | 0.16 | -0.03 – 0.35 | 0.027 |
| Observations | 59 | | |
| R^2^ / R^2^ adjusted | 0.194 / 0.027 | | |

Reference categories are: *Cafes, **IMD quintile 1, ***Liverpool
Negative values indicate a decreased relative difference

**Supplementary Material 5: exploring absolute percentage difference as opposed to relative mean difference**

**Supplementary Table 3: Linear model exploring predictors of absolute percentage difference between reported and measured kcal**

|  | **Absolute percent difference** | | |
| --- | --- | --- | --- |
| *Predictors* | *Estimates* | *99% CI* | *p* |
| (Intercept) | 0.25 | 0.06 – 0.44 | 0.001 |
| **Pubs** | **0.24** | **0.07 – 0.42** | **<0.001** |
| Fast food | 0.07 | -0.10 – 0.24 | 0.264 |
| Restaurant | 0.14 | -0.03 – 0.31 | 0.033 |
| Entertainment | 0.16 | -0.09 – 0.40 | 0.095 |
| Main | -0.03 | -0.20 – 0.13 | 0.625 |
| Dessert | -0.08 | -0.23 – 0.06 | 0.132 |
| Popular | -0.03 | -0.19 – 0.12 | 0.591 |
| Drink | 0.02 | -0.28 – 0.33 | 0.852 |
| IMD quintile [2] | -0.04 | -0.20 – 0.12 | 0.545 |
| IMD quintile [3] | 0.04 | -0.08 – 0.15 | 0.408 |
| IMD quintile [4] | 0.01 | -0.16 – 0.19 | 0.849 |
| IMD quintile [5] | 0.09 | -0.29 – 0.48 | 0.542 |
| Outlet location [MK] | 0.05 | -0.05 – 0.15 | 0.214 |
| **Total energy kcal** | **-0.00** | **-0.00 – 0.00** | **<0.001** |
| Observations | 236 | | |
| R^2^ / R^2^ adjusted | 0.148 / 0.094 | | |

Reference categories are: *Cafes, ** Starters/sides, ***IMD quintile 1, ****Liverpool

1. Finlay A, Polden M, Putra IGNE, Thorp P, Jones A Dr, Adams J, et al. How accurate are calorie labels in the out-of-home food sector in England? 2024. osf.io/8tfu4.
